# The COVID-19 pandemic and temporal change in metabolic risk factors for cardiovascular disease: a natural experiment within the HELIUS study

**DOI:** 10.1101/2021.11.25.21266856

**Authors:** Bryn Hummel, Mara A Yerkes, Ralf E Harskamp, Henrike Galenkamp, Anton E Kunst, Anja Lok, Irene GM van Valkengoed

## Abstract

**Objective:** We studied the association between the coronavirus disease 2019 (COVID-19) pandemic, including the restrictive measures, and metabolic risk factors for cardiovascular disease (CVD) in women and men. Next, we analysed whether changes in these metabolic risk factors were mediated by psychological and behavioural mechanisms.

**Design:** In this natural experiment, we assessed changes from baseline in metabolic CVD risk factors in the exposed group (whose follow-up measurements were taken during the pandemic), and compared these to the changes in the control group (whose follow-up measurements were taken before the pandemic).

**Participants:** This study used data from 6962 participants from six different ethnic groups (Dutch, South-Asian Surinamese, African Surinamese, Ghanaian, Turkish and Moroccan) of the HELIUS study, based in Amsterdam, the Netherlands. We included women and men without prior CVD, who participated in both the baseline (2011-2015) and follow-up measurements (2019-2021).

**Outcome measures:** Changes between baseline and follow-up measurements in six metabolic CVD risk factors were calculated for systolic and diastolic blood pressure (SBP, DBP), total cholesterol (TC), fasting plasma glucose (FPG), haemoglobin A1c (HbA1c), and estimated glomerular filtration rate (eGFR).

**Results:** The exposed group experienced somewhat less favourable changes over time in SBP, DBP and FPG (the latter only in women) than the control group, while temporal changes in HbA1c and eGFR were more favourable among the control group. For instance, SBP was 1.119 mmHg [0.046, 2.193] higher in exposed than non-exposed women, and 1.380 [0.288, 2.471] in men. Changes in SBP and DBP were partially mediated by changes in behavioural factors, most notably BMI and alcohol consumption.

**Conclusions:** The COVID-19 pandemic, including the restrictive lockdown measures, is associated with a deterioration of several CVD risk factors in women and men. These findings may aid in decision making concerning the management of and the recovery following the pandemic.

**Article Summary:** *Strengths and limitations of this study:*

- The COVID19 pandemic lockdown measures led to a pause in the data collection for the prospective, population-based HELIUS study, which shaped a natural experiment.
- Natural experiments, as quasi-experimental designs, are generally considered stronger than cross-sectional studies.
- Through inverse-probability weighting, this study aimed to account for baseline differences between the control and exposed group.
- We could not adjust for differences in follow-up time that occurred as a result of the restrictive measures, which may have affected estimates of variables that change with age.
- The effects of certain mediators may be underestimated, as the data available for defining these variables were largely based on self-reports.

## Introduction

Cardiovascular disease (CVD) is a major cause of morbidity and mortality worldwide [1], and the coronavirus disease 2019 (COVID-19) pandemic is speculated to cause a surge in CVD [2]. Globally, the COVID-19 pandemic has disrupted life for most people, with lockdowns, social distancing and other measures implemented to reduce the spread of the virus. In addition to the direct effect of the severe acute respiratory syndrome coronavirus 2 (SARS-CoV-2) on CVD [3], the restrictive measures might have led to changes in behavioural and psychosocial factors that could negatively affect metabolic CVD risk factors. Understanding how the current pandemic affects cardiometabolic health could lead to better prevention and monitoring of high-risk groups, and may aid decision making surrounding the management of the current and future virus outbreaks.

The impact of pandemics on metabolic CVD risk factors has not been examined. However, major disruptions of the social or physical environment, e.g., after natural disasters, may affect cardiometabolic health. Natural disasters have been linked to deteriorations in metabolic CVD risk factors such as hypertension (HT), Diabetes Mellitus type 2 (DM) and hypercholesterolemia [4-9].

Besides sex differences in COVID-19, the restrictive measures are known to affect women and men differently socially, behaviourally, and psychologically, for example concerning women’s and men’s work, caregiving, psychological distress and health behaviour, such as diets and exercise [10-18]. These differences might translate into sex differences in the effect of the pandemic on metabolic CVD risk factors. Findings on sex differences in response to natural disasters are mixed, with some studies reporting no differences [4], better [5] or worse [7] health outcomes for women than men. Whether these results can be extrapolated to sex differences in the effect of the current pandemic on metabolic CVD risk factors is yet to be empirically determined.

We carried out a natural experiment on the effect of the pandemic on temporal change in six metabolic CVD risk factors [19]: systolic and diastolic blood pressure (SBP, DBP), total cholesterol (TC), fasting plasma glucose (FPG), Haemoglobin A1c (HbA1c), and estimated glomerular filtration rate (eGFR). We determined whether the effect of the pandemic on metabolic CVD risk factors differed between women and men aged 18-70 without prior CVD. Second, we explored to what extent observed differences were mediated by depressive symptoms, negative life events (NLEs) and health behaviour.

## Methods

### HELIUS

Our study is nested within the population-based Healthy Life in an Urban Setting (HELIUS) study, a multi-ethnic cohort study in Amsterdam, the Netherlands [20]. Baseline data were collected between 2011 and 2015 among 24,789 Dutch, South-Asian Surinamese, African Surinamese, Ghanaian, Moroccan, and Turkish origin women and men aged 18-70 years living in Amsterdam. Potential participants were sampled with a simple random sampling method from the municipality registry, after stratification by ethnicity as defined by registered country of birth [21]. The second wave of data collection among participants started in May 2019 and is currently ongoing. Data were obtained by questionnaire and physical examinations (including biological samples).

### Research Ethics Approval

The HELIUS study has been approved by the AMC Ethical Review Board. All participants provided written informed consent.

### Study design

We performed a natural experiment, comparing participants exposed to the pandemic and the restrictive measures during follow-up measurements to those not exposed. This was possible because follow-up data collection was done partially before and partially after the first lockdown (Figure 1). We can thus distinguish between a control group that was examined between the start of data collection on 15 May 2019 until the pause of data collection on 14 March 2020, when the Netherlands entered into its first lockdown [10], and an exposed group whose data were collected when data collection was resumed after the first lockdown ended on 7 July 2020, and 30 December 2020. The exposed group was examined in a period with increasing infections and corresponding restrictive measures. For a more detailed description of the Dutch lockdown measures, please refer to Yerkes et al., 2020 [10].

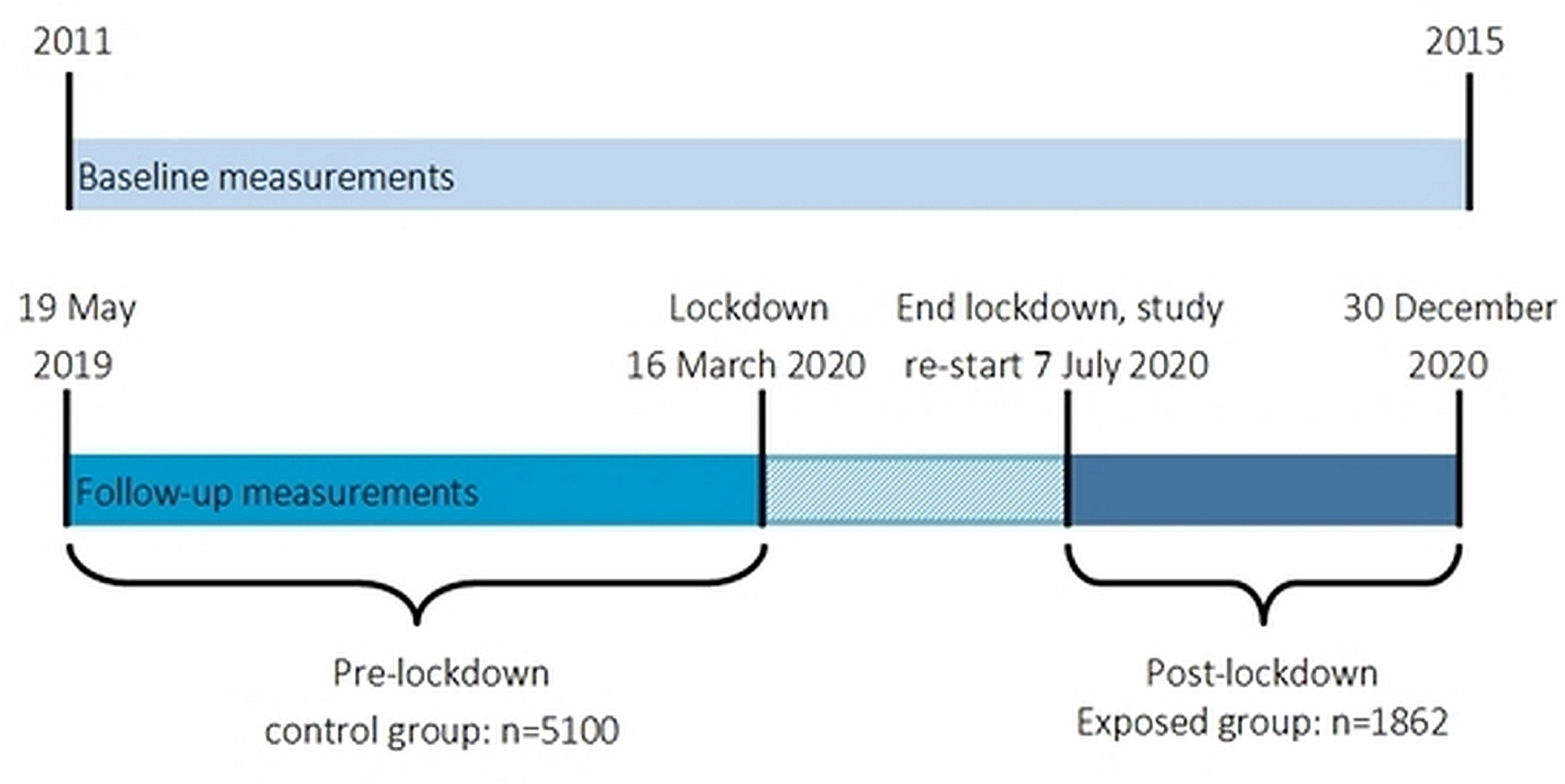

People who participated in both the baseline and follow-up measurement were included (N=8,324). First, those with unknown (N=21), Javanese (N=99) or other Surinamese ethnicity (N=118) were excluded because separate analyses in these groups were not possible due to low power. Next, those with prior CVD (N=1035) or missing data on CVD (n=89) were excluded. The final sample consisted of 6962 participants, 5100 in the control group and 1862 in the exposed group. Cases with missing values were excluded from analyses.

### Metabolic CVD risk factors

We calculated changes from baseline in traditional metabolic CVD risk factors available for both waves, specifically SBP, DBP, HbA1c, FPG, TC, and eGFR [19], by subtracting baseline values from follow-up measurements. Data collection protocols and methods were similar at both time points: SBP and DBP were measured in duplicate on the participants’ left arm using an automated digital blood pressure (BP) device after the participant had been sitting for five minutes. To determine FPG, HbA1c, TC and serum creatinine, fasting blood samples were drawn after an overnight fast. FPG was determined in plasma samples by using enzymatic spectrophotometric UV method, using hexokinase as primary enzyme (Roche Diagnostics), and HbA1c was determined in whole blood samples through HPLC technology (Tosoh). Serum creatinine was determined using a kinetic colorimetric spectrophotometric isotope dilution mass spectrometry-calibrated method (Roche Diagnostics), and eGFR was calculated using the CKDEPI (CKD epidemiology collaboration) creatinine equation [22].

### Covariates

Socio-demographic data were assessed by questionnaire. Ethnicity was defined by the participant’s and their parents’ country of birth [21]: a person was defined as belonging to one of the included minority groups if (1) he/she was born outside the Netherlands with at least one parent born outside the Netherlands (first generation) or (2) he/she was born in the Netherlands but both parents were born outside the Netherlands (second generation). For the Dutch sample, people born in the Netherlands whose parents were also born in the Netherlands, were invited. Surinamese participants were further classified according to self-reported ethnic origin into ‘African’, ‘South-Asian’ or ‘other’. We used highest attained educational level, labour market participation and occupational level as a proxies for socioeconomic status (SES). Educational level was categorized into three categories: lower (no education, elementary education or lower vocational or lower secondary education), intermediate (intermediate vocational or intermediate or higher secondary education), or high (higher vocational education or university). Labour market participation consisted of four categories: employed, not in employment (including retirees, students, homemakers), unemployed and/or social benefit recipients, and occupationally disabled. Occupational level was classified according to the Dutch Standard Occupational Classification system, which provides an extensive systematic list of all professions in the Dutch system. Occupational level consisted of five categories, based on job title and job description [23], and consisted of five levels: elementary, lower, intermediate, higher, and scientific occupations.

### Mediators

Changes in depressive symptoms was calculated by subtracting baseline from follow-up values. Depressive symptoms were assessed via the Patient Health Questionnaire (PHQ-9), a validated instrument [24] measuring depressive mood in the past two weeks. Change in BMI was calculated by subtracting baseline BMI from follow-up BMI, with weight measured in light clothing and height measured without shoes [20]. Self-reported smoking status, alcohol use and NLE were assessed via questionnaire. Smoking status measured whether people are current smokers, have never smoked or have previously smoked, which was recoded into those who quit or started smoking between baseline and follow-up, and no change. Alcohol use consisted of the categories low, intermediate and high, which was recoded into decreased, equal or increased alcohol use between baseline and follow-up. NLEs at follow-up assessed whether people reported recently experiencing a NLE, such as illness, death of a loved one or other adverse experiences, recoded into yes or no.

### Data analysis

Socio-demographic variables and mediators were presented as means [standard deviations (SD)], or frequencies [percentages], by control or exposed group and sex.

We performed linear regression analyses, stratified by sex, with change in metabolic risk factors as dependent variables, and exposure to the lockdown as the main predictor. Possible selection bias and the effect of phasing of the HELIUS data collection (i.e., for organizational reasons different neighbourhoods in Amsterdam were invited sequentially) could lead to baseline differences between the control and exposed group due to social and ethnic characteristics of neighbourhoods. To account for these differences, we used inverse probability weighting (IPW) on baseline values of the metabolic CVD risk factors and sociodemographic factors [25]. This means weights are created for the likelihood of being exposed to the pandemic based on baseline factors, to balance the control and exposed group.

Because of the effect of the pandemic on behavioural and psychosocial factors, we explored the mediating effect of behavioural and psychosocial factors on the association between exposure to the pandemic and change in metabolic risk factors for which a difference between the control and exposed group was found. Specifically, we described change in smoking status, alcohol use, BMI and depressive symptoms, and the presence of recent NLEs [2,16,17,26], and compared the model without mediators to the model with all mediators to measure whether these psychological and behavioural mechanisms contributed to the observed differences between the control and exposed group. Next, we added the mediators separately to see which factors contributed most to these changes. Decreases of ≥10% in the beta’s were considered to be indicative of a mediation effect. For eGFR, we additionally explored to what extent changes in SBP and DBP explained observed differences.

As medication use or seasonal effects might affect comparisons, we repeated the main analyses, first, after excluding participants who were receiving HT and/or DM medication at baseline and, second, after excluding participants measured between January and June from the control group to ensure comparisons across similar time periods [27].

Additionally, we verified if differences were present across educational levels (as a proxy of socioeconomic status; SES) and ethnic groups, given that the pandemic may differently affect various groups within society [10,11,14,28].

Finally, we conducted a supplemental analysis to explore to what degree observed effects varied across time. We distinguished between two time periods in the exposed group: the ‘early post-lockdown period’ between data collection resumption until 21 September 2020, representing a period with limited measures and a low infection rate after the first wave subsided, and 21 September until 30 December 2020, the ‘late post-lockdown period’, with implementation of new measures and increasing infection rates (the second wave [28]).

The analyses were conducted in R studio 4.0.3, with statistical significance determined at p-values <.05.

## Results

### Sample characteristics

56.2% of participants was female, the mean age was 45.8 (SD=12.5) at baseline and 52.0 (SD=12.6) at follow-up, with an average follow-up time of 6 years and 2 months (Table 1). The exposed group was significantly younger, more often higher educated and more often of Dutch origin than the control group. All baseline CVD risk factors, except TC, were more favourable among the exposed group.

**Table 1.**
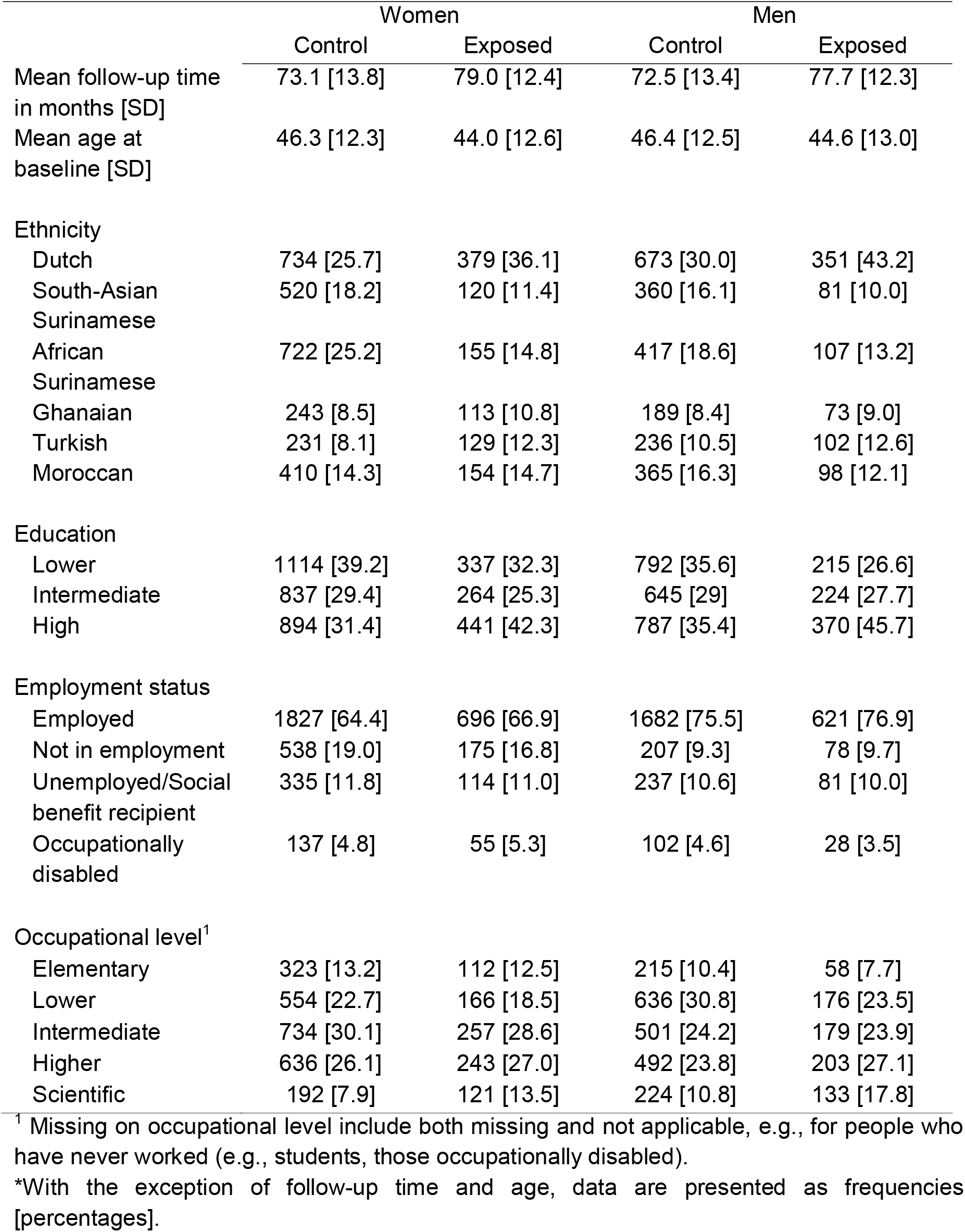
Sociodemographic characteristics by sex and quasi-experimental group*.

### Differences control and exposed group

We found small, yet statistically significant differences in crude mean change in metabolic CVD risk factors between the control and exposed group in women and men (Supplemental Table 1). SBP, DBP, and FPG increased more, and eGFR decreased less, in the exposed group than the control group. Change in TC was similar between groups, whereas changes in HbA1c decreased more in the exposed group.

In analyses weighted on baseline characteristics, these differences remained (Table 2, Figure 2). In the exposed group, women appeared to have slightly greater deteriorations in DBP and FPG, and smaller improvements in HbA1c compared to men, while men appeared to experience larger deteriorations in SBP and slightly smaller improvements in eGFR. These sex differences, however, were not statistically significant.

**Table 2.** Linear regressions on the association between exposure to the pandemic, including lockdown measures, and temporal change in metabolic risk factors, by sex*.

|  | Women |  | Men |  | Interaction |  |
| --- | --- | --- | --- | --- | --- | --- |
|  | B [95% CI] | p-value | B [95% CI] | p-value | B [95% CI] | p-value |
| ΔSBP | 1.119 [0.046, 2.193] | .041 | 1.380 [0.288, 2.471] | .013 | -0.699 [-2.249, 0.851] | .377 |
| ΔDBP | 0.848 [0.232, 1.463] | .007 | 0.797 [0.107, 1.488] | .024 | -0.044 [-0.969, 0.882] | .927 |
| ΔTC | -0.054 [-0.117, 0.009] | .091 | -0.005 [-0.074, 0.064] | .886 | -0.052 [-0.146, 0.042] | .276 |
| ΔFPG | 0.121 [0.056, 0.185] | .000 | 0.022 [-0.066, 0.109] | .626 | 0.073 [-0.036, 0.181] | .189 |
| ΔHbA1c | -0.645 [-0.974, -0.316] | .000 | -0.835 [-1.370, -0.300] | .002 | 0.115 [-0.493, 0.722] | .712 |
| ΔeGFR | 1.062 [0.387, 1.737] | .002 | 1.041 [0.322, 1.760] | .005 | -0.023 [-1.008, 0.961] | .963 |

\*SBP, systolic blood pressure; DBP, diastolic blood pressure; TC, total cholesterol; FPG, fasting plasma glucose; HbA1c, haemoglobin A1c; eGFR, estimated glomerular filtration rate; CI, confidence intervals. The analyses were weighted on baseline measurements, age at baseline, educational level, ethnicity, occupational level and labour market participation. The interaction column tests the interaction between exposure to the pandemic and lockdown and sex.

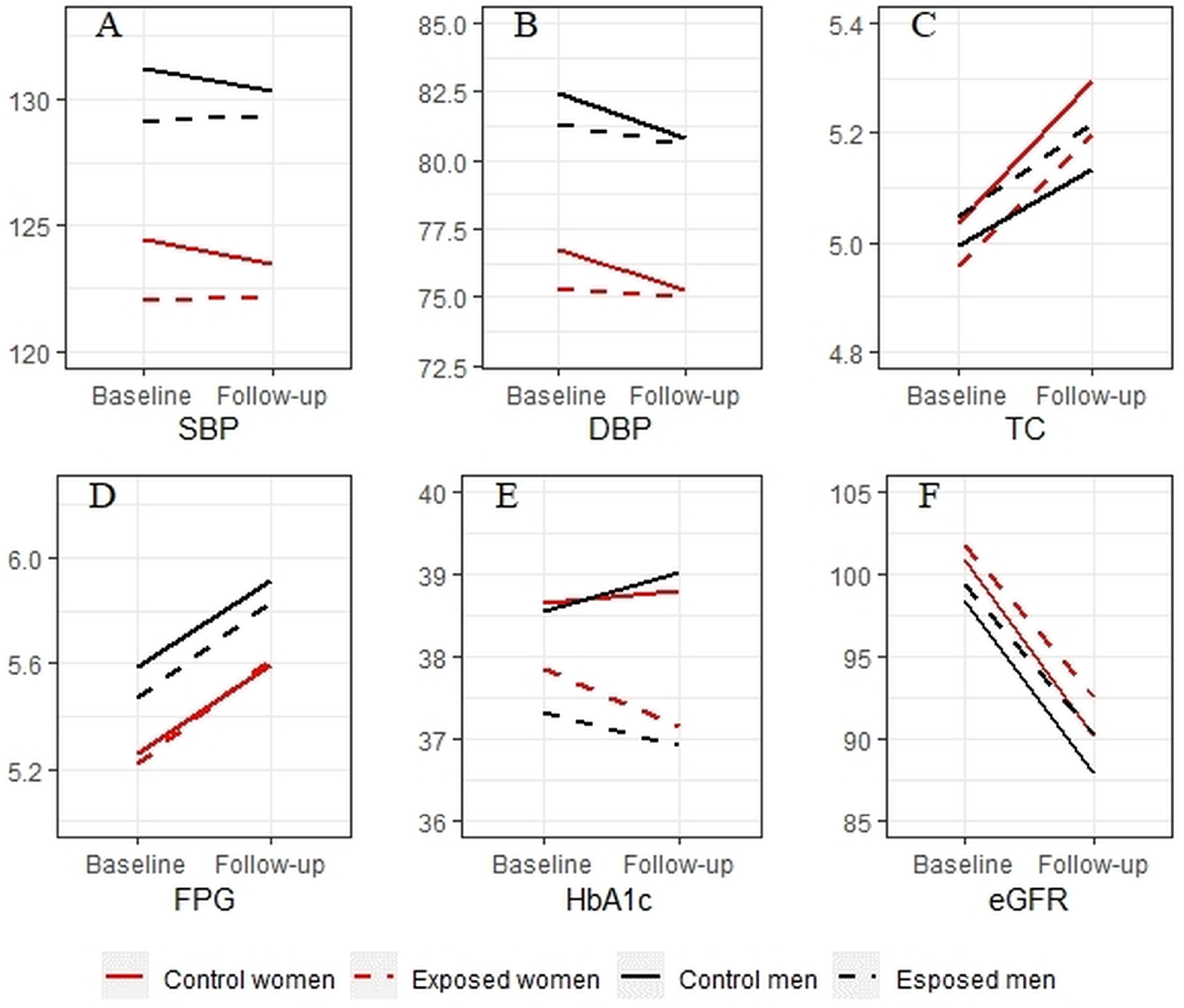

### Mediation

We found significant differences in several potential mediators between the control and exposed group, such as a larger increase in BMI in the exposed group than the control group (Supplemental Table 6). Addition of these mediators partially explained observed differences in several risk factors between the control and exposed group. For instance, in the model with all mediators, the beta’s of exposure to the pandemic on DBP decreased with 50.8% for women and 90% for men (Table 3). Changes in BMI and alcohol use, and smoking status for women, were associated with the largest changes in beta’s for SBP, DBP and FPG (in men), but not HbA1c. Change in alcohol use, but not other mediators, nor SBP and DBP, partially mediated the relationship between exposure to the pandemic and change in eGFR (−11.8% among men compared -35.3% among women).

**Table 3.** Linear regression on the association between exposure to the pandemic, including lockdown measures, and temporal change in metabolic risk factors, adjusting for mediators*.

|  | Women |  |  | Men |  |  |
| --- | --- | --- | --- | --- | --- | --- |
|  | B [95% CI] | p-value | %Δ in B | B [95% CI] | p-value | %Δ in B |
| ΔSBP main analysis | 1.119 [0.046, 2.193] | .041 |  | 1.380 [0.288, 2.471] | .013 |  |
| + All mediators | 0.428 [-0.800, 1.655] | .495 | - 69.1% | 0.343 [-0.883, 1.568] | .583 | - 75.1% |
| + Alcohol | 0.962 [-0.277, 2.202] | .128 | - 14.0% | 0.762 [-0.489, 2.013] | .233 | - 44.8% |
| + Smoking | 0.978 [-0.098, 2.053] | .075 | - 12.6% | 1.357 [0.264, 2.450] | .015 | -1.7% |
| + BMI | 0.787 [-0.270, 1.844] | .145 | - 29.7% | 1.021 [-0.048, 2.089] | .061 | - 26.0% |
| + PHQ-9 | 1.072 [-0.005, 2.150] | .051 | -4.2% | 1.320 [0.225, 2.415] | .018 | -4.3% |
| + NLE | 1.090 [0.015, 2.166] | .047 | -2.6% | 1.380 [0.288, 2.471] | .013 | 0.0% |
| ΔDBP main analysis | 0.848 [0.232, 1.463] | .007 |  | 0.797 [0.107, 1.488] | .024 |  |
| + All mediators | 0.417 [-0.282, 1.117] | .243 | - 50.8% | 0.080 [-0.683, 0.843] | .837 | - 90.0% |
| + Alcohol | 0.712 [-0.001, 1.426] | .050 | - 16.0% | 0.432 [-0.359, 1.223] | .284 | - 45.8% |
| + Smoking | 0.777 [0.161, 1.393] | .013 | -8.4% | 0.758 [0.067, 1.450] | .032 | -4.9% |
| + BMI | 0.617 [0.017, 1.217] | .044 | - 27.2% | 0.515 [-0.153, 1.182] | .131 | - 35.4% |
| + PHQ-9 | 0.838 [0.220, 1.456] | .008 | -1.2% | 0.725 [0.032, 1.418] | .040 | -9.0% |
| + NLE | 0.845 [0.229, 1.461] | .007 | -0.4% | 0.793 [0.103, 1.484] | .024 | -0.5% |
| ΔFPG main analysis | 0.121 [0.056, 0.185] | .000 |  | 0.022 [-0.066, 0.109] | .626 |  |
| + All mediators | 0.137 [0.065, 0.209] | .000 | +13.2 % | 0.006 [-0.095, 0.107] | .908 | - 72.7% |
| + Alcohol | 0.141 [0.066, 0.215] | .000 | +16.5 % | 0.012 [-0.089, 0.112] | .820 | - 45.5% |
| + Smoking | 0.120 [0.055, 0.185] | .000 | -0.8% | 0.027 [-0.061, 0.114] | .554 | +22.7 % |
| + BMI | 0.110 [0.045, 0.174] | .001 | -9.0% | 0.013 [-0.075, 0.100] | .776 | - 40.9% |
| + PHQ-9 | 0.127 [0.064, 0.190] | .000 | +5.0% | 0.019 [-0.070, 0.107] | .679 | - 13.6% |
| + NLE | 0.121 [0.056, 0.186] | .000 | 0.0% | 0.022 [-0.066, 0.109] | .628 | 0.0% |
| ΔHbA1c main analysis | -0.645 [-0.974, -0.316] | .000 |  | -0.835 [-1.370, -0.300] | .002 |  |
| + All mediators | -0.678 [-1.053, -0.304] | .000 | +5.1% | -0.821 [-1.434, -0.209] | .009 | -1.7% |
| + Alcohol | -0.625 [-1.008, -0.242] | .001 | -3.1% | -0.808 [-1.419, -0.197] | .010 | -3.2% |
| + Smoking | -0.641 [-0.972, -0.311] | .000 | -0.6% | -0.795 [-1.330, -0.260] | .004 | -4.8% |
| + BMI | -0.731 [-1.056, -0.406] | .000 | +13.3% | -0.892 [-1.427, -0.357] | .001 | +6.8% |
| + PHQ-9 | -0.620 [-0.946, -0.295] | .000 | -3.9% | -0.861 [-1.399, -0.323] | .002 | +3.1% |
| + NLE | -0.647 [-0.976, -0.317] | .000 | +3.1% | -0.838 [-1.373, -0.304] | .002 | +3.6% |
| ΔeGFR main analysis | 1.062 [0.387, 1.737] | .002 |  | 1.041 [0.322, 1.760] | .005 |  |
| + All mediators | 0.690 [-0.088, 1.468] | .082 | -35.0% | 1.121 [0.303, 1.939] | .007 | +7.7% |
| + Alcohol | 0.687 [-0.088, 1.462] | .082 | -35.3% | 0.918 [0.096, 1.739] | .029 | -11.8% |
| + Smoking | 1.065 [0.387, 1.743] | .002 | +0.3% | 1.070 [0.351, 1.788] | .004 | +2.8% |
| + BMI | 1.110 [0.435, 1.785] | .001 | +4.5% | 1.193 [0.480, 1.907] | .001 | +14.6% |
| + PHQ-9 | 0.997 [0.320, 1.673] | .004 | -6.1% | 1.096 [0.374, 1.818] | .003 | +5.3% |
| + NLE | 1.073 [0.399, 1.747] | .002 | +1.0% | 1.041 [0.323, 1.760] | .005 | 0.0% |
| + SBP and DBP | 1.025 [0.350, 1.700] | .003 | -3.5% | 1.015 [0.295, 1.735] | .006 | -2.5% |

\*SBP, systolic blood pressure; DBP, diastolic blood pressure; TC, total cholesterol; FPG, fasting plasma glucose; HbA1c, haemoglobin A1c; eGFR, estimated glomerular filtration rate; BMI, body mass index; PHQ, patient health questionnaire; NLE, negative life event; CI, confidence intervals. Mediation for TC is not reported because TC did not differ between the control and exposed group in the main analyses. The analyses were weighted on baseline measurements, age at baseline, educational level, ethnicity, occupational level and labour market participation.

### Supplemental analyses

Analyses excluding participants receiving HT and/or DM treatment and the analyses for seasonal effects did not change our interpretation of these findings (Supplemental Tables 2-3). Moreover, analyses stratified by SES and ethnicity, except for African Surinamese women and Turkish men, confirmed the association between the pandemic on SBP, DBP and FPG, but not other measures (Supplemental Tables 4-5). However, groups were too small and confidence intervals too wide to assess whether differences within and between women and men varied by SES or ethnicity.

Finally, the analysis of time periods revealed that in both women and men, the associations between exposure to the pandemic and SBP, DBP, FPG and TC (in men), were larger in the late post-lockdown period compared to the early post-lockdown period (Supplemental Table 7).

## Discussion

Women and men in the exposed group experienced slightly higher increases in SBP, DBP and FPG than the control group. Change in TC did not differ between groups, while the exposed group experienced slightly more favourable changes in HbA1c and eGFR relative to the control group. Patterns of differences did not vary significantly between women and men, and were observed across most ethnic and SES groups. Particularly in men, but also in women, changes in SBP, DBP, FPG and eGFR may have been partially mediated by behavioural factors, most notably changes in BMI and alcohol use. Psychosocial factors did not contribute substantially.

While natural experiments are generally considered strong [29], some limitations persist. First, response bias may have occurred [20] affecting generalizability to the general population. Moreover, random assignment to the control or exposed group was not possible, resulting in baseline differences between these groups. This may be due to phasing of the study, or selective drop-out among exposed participants. For instance, if only healthy participants were comfortable with being examined during the pandemic, this would result in a healthy subset of participants. We attempted to account for these differences through IPW, yet, this method does not account for potentially relevant unmeasured confounders. Despite this limitation, we chose IPW as our method to adjust for baseline differences and possible selective drop-out as it uses the entire sample, whereas propensity score matching would have led to a loss of cases, thereby negatively affecting our power [25].

Moreover, we could not adjust for differences in follow-up time, as this is a direct result of the restrictive measures. Yet, this may have affected variables that change with age, e.g. eGFR and SBP. On average, SBP increases with 7 mmHg every ten years [30] and eGFR decreases with 1 mL/min/year rate [31]. Given that the difference in follow-up time between the control and exposed group is only several months, this is thus unlikely to completely explain our findings.

Furthermore, the effect of the mediators may have been underestimated, as measures characterizing change in behaviour may have been suboptimal, for example, only change in smoking status was available instead of amount and products smoked. Moreover, while we used the most common method of assessing these variables [20], some mediators were self-report data, which may be less accurate compared to other measurement methods [32].

Despite these limitations, we found significant differences in several metabolic CVD risk factors between the control and exposed group, suggesting a potential effect of the pandemic on future CVD risk in both women and men. The magnitude of the associations appears modest, since an average increase of e.g. 1.38 mmHg in SBP may not drastically alter an individual’s CVD risk [33]. Yet, observed effects could potentially be problematic at the population level. Indeed, associations were found across SES- and ethnic subgroups, and density plots of several risk factors control or exposed group revealed that the difference reflects a slight shift in the entire distribution of risk factors (data not shown). Thus, in line with Rose’s prevention paradox, this could imply a need for intervention strategies aimed at the total population rather than exclusively high-risk individuals [34].

In particular, our findings suggest the pandemic negatively affected BP and glycaemia. Elevated BP due to the pandemic is in line with prior research on social disruption following natural disasters [4,6-8]. Moreover, in line with prior research [8], we found evidence that the pandemic led to increased glycaemia, in particular FPG among women. Findings concerning HbA1c seem contradictory, as the exposed group had slightly more favourable changes in HbA1c than the control group, despite the less favourable changes in FPG. This could be due to the inclusion of people with DM in our study, as studies have described better DM self-management during the lockdown due to improved medication compliance and adherence to dietary recommendations [15]. However, analyses excluding people receiving DM treatment show this is unlikely to explain these findings. Alternatively, HbA1c as a long-term measure of glycaemia may be less accurate in capturing the acute effects of the pandemic [35], which is supported by our finding that HbA1c values are worse in the late post-lockdown period.

We found no unfavourable association between exposure to the pandemic and other risk factors. Previous studies reported increased cholesterol following large disruptive events [5,6,9]. We found no effect on TC, despite the larger increase in BMI in the exposed group, which is evidently associated with TC [36]. We speculate this may be due to the specific underlying dietary- and lifestyle patterns in our population, which may associate more strongly with other measures of cholesterol than TC, such as HDL, LDL or triglycerides [36]. Moreover, contrary to our expectations, the exposed group experienced a smaller decline in eGFR than the control group. This decline was not likely due to hyperfiltration, as changes in SBP and DBP did not mediate the association between exposure to the pandemic and change in eGFR. Neither was this likely due to COVID-19 infections, as the association between the SARS-CoV-2 virus and acute kidney injury is small [37], and because the association was not larger in the ethnic group with the highest infection rate, Ghanaians [28], which would be expected if this effect was caused by COVID-19 infections. A more feasible explanation seems an overestimation of the eGFR, as creatinine levels at follow-up may have been influenced by changes in muscle mass, for instance due to decreased exercise during the lockdown [38].

The association between exposure to the pandemic and BP and glycaemia seem to be partially mediated by behavioural factors, in particular BMI and alcohol use, two well-known CVD risk factors. These observed differences are in line with studies on health behaviour during the pandemic [12,13,15-17] and changes reported following natural disasters [6,39]. Strategies to promote healthy diets and exercise, reduce alcohol use and smoking could potentially be used in the recovery following the pandemic [40]. While several studies reported increased mental distress as a result of the pandemic [11,18], we found no effect of recent NLEs or change in depressive symptoms. However, given the continuation of the pandemic and the restrictive measures, long term implications of mental distress due to the pandemic are to be evaluated in prospective research. The mediating effects of other social and behavioural factors not assessed in this study, warrant further inquiry.

We found no evidence supporting sex differences in the effects of the pandemic, despite a body of literature reporting sex differences in changes in health behaviour due to the pandemic. Studies have reported unhealthier diets [14] and more psychological problems [11,18] among women, and worse sleeping patterns, decreased exercise [12], higher alcohol consumption [16] and higher risk of DM [13] among men during the pandemic. Thus, while the pandemic may have affected women and men differently socially, behaviourally and psychologically, this might not have translated into sex differences in the effect of the pandemic on CVD risk factors in the short term. Future research should determine whether this trend continues over the duration of the pandemic.

We conclude that the pandemic may have caused a modest deterioration of metabolic CVD risk factors in both women and men across ethnic and socioeconomic groups. These effects seem to be partially mediated by changes in health behaviour, possibly as a result of the restrictive measures during the pandemic. These results suggest a need to prepare for, and mitigate, potential long-term effects, and may be valuable in decision making surrounding the management of – and recovering following – the COVID-19 pandemic.

## Data Availability

The HELIUS data are owned by the Amsterdam University Medical Centers, location AMC in Amsterdam, The Netherlands. Any researcher can request the data by submitting a proposal to the HELIUS Executive Board as outlined at http://www.heliusstudy.nl/en/researchers/collaboration, by. The HELIUS Executive Board will check proposals for compatibility with the general objectives, ethical approvals and informed consent forms of the HELIUS study. There are no other restrictions to obtaining the data and all data requests will be processed in the same manner.

## Declarations

## Acknowledgments

The HELIUS study is conducted by the Amsterdam University Medical Centre, location AMC and the Public Health Service of Amsterdam. Both organizations provided core support for HELIUS. We are most grateful to the participants of the HELIUS study and the management team, research nurses, interviewers, research assistants and other staff who have taken part in gathering the data of this study. We also want to thank Dr. Ehsan Motazedi for critically reviewing our methodology.

## Authors Contributions

Contributors BH and IGMvV contributed to the conception and design of the work. BH drafted the manuscript. IGMvV, MAY, REH, HG, AEK, and AL critically revised the manuscript. All authors gave final approval and agree to be accountable for all aspects of work ensuring integrity and accuracy. The corresponding author attests that all listed authors meet authorship criteria and that no others meeting the criteria have been omitted. All gave final approval and agree to be accountable for all aspects of work ensuring integrity and accuracy.

## Patient and Public Involvement

It was not appropriate or possible to include patients or the public in the design, or conduct, or reporting, or dissemination plans of our research

## References

1. WHO. Cardiovascular diseases (CVDs)2017 2021 Jan 28. Available from: https://www.who.int/news-room/fact-sheets/detail/cardiovascular-diseases-(cvds).

2. Virani SS, Alonso A, Aparicio HJ, Benjamin EJ, Bittencourt MS, Callaway CW, et al. Heart Disease and Stroke Statistics—2021 Update: A report From the American Heart Association. Circulation. 2021:e1–e143.

3. Adu-Amankwaah J, Mprah R, Adekunle AO, Ndzie Noah ML, Adzika GK, Machuki JO, et al. The cardiovascular aspect of COVID-19. Annals of Medicine. 2021;53(1):227–36.

4. Li C, Luo X, Zhang W, Zhou L, Wang HC.Z. QaAn earthquake increases blood pressure among hospitalized patients. Clinical and Experimental Hypertension. 2016;38(6):495–9.

5. Li N, Wang Y, Yu L, Song M, Wang L, Ji C, et al. Long-term effects of earthquake experience of young persons on cardiovascular disease risk factors. Arch Med Sci. 2017;13(1):75–81.

6. Shiba K, Hikichi H, Aida J, Kondi K, Kawachi I. Long-Term Associations between Disaster Experiences and Cardiometabolic Risk: A Natural Experiment From the 2011 Great East Japan Earthquake and Tsunami. American Journal of Epidemiology. 2019;188(6):1109–19.

7. Vuković I, Jovanović N, Kulenović A, Britvić D, Mollica Rf. Women health: Psychological and most prominent somatic problems in 3-year follow-up in Bosnian refugees. International Journal of Social Psychiatry. 2020.

8. Gohardehi F, Seyedin H, Moslehi S. Prevalence Rate of Diabetes and Hypertension in Disaster-Exposed Populations: A Systematic Review and Meta-Analysis. Ethiop J Health Sci. 2020;30(3):439–48.

9. Trevisan M, Jossa F, Farinaro E, Krogh V, Panico S, Giumetti D, et al. Earthquake and coronary heart disease risk factors: a longitudinal study. American Journal of Epidemiology. 1992;135(6):632–7.

10. Yerkes MA, André SCH, Besamusca JW, Kruyen PM, Remery Clhs, van der Zwan R, et al. ‘Intelligent’ lockdown, intelligent effects? Results from a survey on gender (in)equality in paid work, the division of childcare and household work, and quality of life among parents in the Netherlands during the Covid-19 lockdown. PLoSONE. 2020;15(11).

11. Pierce M, Hope H, Ford T, Hatch S, Hotopf M, John A, et al. Mental health before and during the COVID-19 pandemic: a longitudinal probability sample survey of the UK population. Lancet Psychiat. 2020;7(10):883–92.

12. Barrea L, Pugliese G, Framondi L, Di Matteo R, Laudisio D, Savastano S, et al. Does Sars-Cov-2 threated our dreams? Effect of quarantine on sleep quality and body mass index. Journal of Translational Medicine. 2020;18(1):318–29.

13. Ghosal S, Arora B, Dutta K, Ghosh A, Sinha B, Misra A. Increase in the risk of type 2 diabetes during lockdown for the COVID19 pandemic in India: A cohort analysis. Diabetes & metabolic syndrome. 2020;14(5):949–52.

14. Poelman MP, Gillebaart M, Schlinkert C, Dijkstra SC, Derksen E, Mensink F, et al. Eating behavior and food purchases during the COVID-19 lockdown: A cross-sectional study among adults in the Netherlands. Appetite. 2020;157:105002.

15. Rastogi A, Hiteshi P, Bhansali A. Improved glycemic control amongst people with long-standing diabetes during COVID-19 lockdown: a prospective, observational, nested cohort study. Int J Diabetes Dev Ctries. 2020:1–6.

16. Bakaloudi DR, Jeyakumar DT, Jayawardena R, Chourdakis M. The impact of COVID-19 lockdown on snacking habits, fast-food and alcohol consumption: A systematic review of the evidence. Clin Nutr. 2021.

17. Kreutz R, Dobrowolski P, Prejbisz A, Algharably EAEH, Bilo G, Creutzig F, et al. Lifestyle, psychological, socioeconomic and environmental factors and their impact on hypertension during the coronavirus disease 2019 pandemic. Journal of Hypertension. 2021;39(6):1077–89.

18. Vanderlind WM, Rabinovitz BB, Miao IY, Oberlin LE, Bueno-Castellano C, Fridman C, et al. A systematic review of neuropsychological and psychiatric sequalae of COVID-19: implications for treatment. Curr Opin Psychiatry. 2021;34(4):420–33.

19. Ramezankhani A, Azizi F, Hadaegh F, Eskandari F. Sex-specific clustering of metabolic risk factors and their association with incident cardiovascular diseases: A population-based prospective study. Atherosclerosis. 2017;263:249–56.

20. Snijder MB, Galenkamp H, Prins M, Derks EM, Peters RJG, Zwinderman AH, et al. Cohort profile: the Healthy Life in an Urban Setting (HELIUS) study in Amsterdam, The Netherlands. BMJ Open. 2017;7(12):e017873.

21. Stronks K, Kulu-Glasgow I, Agyemang C. The utility of ‘country of birth’ for the classification of ethnic groups in health research: the Dutch experience. Ethnic Health. 2009;14(3):255–69.

22. Levey AS, Stevens LA. Estimating GFR Using the CKD Epidemiology Collaboration (CKD-EPI) Creatinine Equation: More Accurate GFR Estimates, Lower CKD Prevalence Estimates, and Better Risk Predictions. American Journal of Kidney Diseases. 2010;55(4):622–7.

23. Netherlands S. Standaard Beroepenclassificatie 2010. Den Haag: Statistics Netherlands; 2010.

24. Galenkamp H, Stronks K, Snijder MB, Derks EM. Measurement invariance testing of the PHQ-9 in a multi-ethnic population in Europe: the HELIUS study. BMC Psychiatry. 2017;17(1):349.

25. Sheikhy A, Fallahzadeh A, Sadeghian S, Forouzannia K, Bagheri J, Salehi-Omran A, et al. Mid-term outcomes of off-pump versus on-pump coronary artery bypass graft surgery; statistical challenges in comparison. BMC Cardiovasc Disord. 2021;21(1):412.

26. Havranek EP, Mujahid MS, Barr DA, Blair IV, Cohen MS, Cruz-Flores S, et al. Social Determinants of Risk and Outcomes for Cardiovascular Disease A Scientific Statement From the American Heart Association. Circulation. 2015;132(9):873–98.

27. Aubiniere-Robb L, Jeemon P, Hastie CE, Patel RK, McCallum L, Morrison D, et al. Blood pressure response to patterns of weather fluctuations and effect on mortality. Hypertension. 2013;62(1):190–6.

28. Coyer L, Boyd A, Schinkel J, Agyemang C, Galenkamp H, Koopman ADM, et al. SARS-CoV-2 antibody prevalence and determinants of six ethnic groups living in Amsterdam, the Netherlands: 2 a population-based cross-sectional study, June-October 2020. 2021:1–33.

29. Leatherdale ST. Natural experiment methodology for research: a review of how different methods can support real-world research. Int J Soc Res Method. 2019;22(1):19–35.

30. Gurven M, Blackwell AD, Rodríguez DE, Stieglitz J, Kaplan H. Does Blood Pressure Inevitably Rise With Age? Longitudinal Evidence Among Forager-Horticulturalists. Hypertension. 2012;60(1):25–33.

31. Nojima J, Meguro S, Ohkawa N, Furukoshi M, Kawai T, Itoh H. One-year eGFR decline rate is a good predictor of prognosis of renal failure in patients with type 2 diabetes. P Jpn Acad B-Phys. 2017;93(9):746–54.

32. Hebert JR. Social Desirability Trait: Biaser or Driver of Self-Reported Dietary Intake? J Acad Nutr Diet. 2016;116(12):1895–8.

33. Conroy RM, Pyorala K, Fitzgerald AP, Sans S, Menotti A, De Backer G, et al. Estimation of ten-year risk of fatal cardiovascular disease in Europe: the SCORE project. European Heart Journal. 2003;24(11):987–1003.

34. Raza SA, Salemi JL, Zoorob RJ. Historical perspectives on prevention paradox: When the population moves as a whole. Journal of Family Medical Primary Care. 2018;7(6):1163–5.

35. Rohlfing CL, Wiedmeyer HM, Little RR, England JD, Tennill A, Goldstein DE. Defining the Relationship Between Plasma Glucose and HbA1c: analysis of glucose profiles and HbA1c in the Diabetes Control and Complications Trial. Diabetes Care. 2002;25(2):275–8.

36. Muga MA, Owili PO, Hsu CY, Chao JC. Association of lifestyle factors with blood lipids and inflammation in adults aged 40 years and above: a population-based cross-sectional study in Taiwan. BMC Public Health. 2019;19(1):1346.

37. Singh J, Malik P, Patel N, Pothuru S, Israni A, Chakinala RC, et al. Kidney disease and COVID-19 disease severity-systematic review and meta-analysis. Clin Exp Med. 2021.

38. Baxmann AC, Ahmed MS, Marques NC, Menon VB, Pereira AB, Kirsztajn GM, et al. Influence of muscle mass and physical activity on serum and urinary creatinine and serum cystatin C. Clin J Am Soc Nephrol. 2008;3(2):348–54.

39. Terayama T, Shigemura J, Kobayashi Y, Kurosawa M, Nagamine M, Toda H, et al. Mental health consequences for survivors of the 2011 Fukushima nuclear disaster: a systematic review. Part 2: emotional and behavioral consequences. CNS Spectr. 2021;26(1):30–42.

40. Mattioli AV, Ballerini Puviani M, Nasi M, Farinetti A. COVID-19 pandemic: the effects of quarantine on cardiovascular risk. Eur J Clin Nutr. 2020;74(6):852–5.

