## Supplemental for "The COVID-19 pandemic and temporal change in metabolic risk factors for cardiovascular disease: a natural experiment within the HELIUS study"

SUPPLEMENTAL TABLES

**Supplemental Table 1. Metabolic CVD risk factors at baseline, follow-up and change over time by quasi-experimental group and sex**

|  | Women | | Men | |
| --- | --- | --- | --- | --- |
|  | Control | Exposed | Control | Exposed |
| SBP |  |  |  |  |
| SBP baseline | 124.5 [17.52] | 122.0 [17.55] | 131.2 [15.42] | 129.1 [15.50] |
| SBP follow-up | 123.5 [18.23] | 122.1 [19.44] | 130.3 [16.26] | 129.3 [16.20] |
| ∆SBP | -1.00 [14.04] | 0.07 [14.76] | -0.88 [13.26] | 0.22 [14.05] |
| DBP |  |  |  |  |
| DBP baseline | 76.76 [10.08] | 75.34 [10.03] | 82.44 [9.78] | 81.29 [10.24] |
| DBP follow-up | 75.26 [9.94] | 75.03 [10.17] | 80.80 [9.90] | 80.60 [9.98] |
| ∆DBP | -1.50 [7.97] | -0.31 [8.46] | -1.67 [8.10] | -0.68 [9.0] |
| TC |  |  |  |  |
| TC baseline | 5.04 [1.02] | 4.96 [0.98] | 4.99 [1.01] | 5.05 [1.04] |
| TC follow-up | 5.30 [1.06] | 5.21 [1.10] | 5.13 [1.06] | 5.21 [1.11] |
| ∆TC | 0.26 [0.88] | 0.24 [0.83] | 0.14 [0.88] | 0.17 [0.89] |
| FPG |  |  |  |  |
| FPG baseline | 5.26 [0.93] | 5.22 [1.09] | 5.59 [0.97] | 5.47 [0.88] |
| FPG follow-up | 5.60 [1.20] | 5.62 [1.06] | 5.92 [1.51] | 5.83 [1.15] |
| ∆FPG | 0.34 [0.86] | 0.41 [1.04] | 0.33 [1.18] | 0.36 [0.95] |
| HbA1c |  |  |  |  |
| HbA1c baseline | 38.64 [7.01] | 37.86 [7.47] | 38.56 [7.54] | 37.34 [6.73] |
| HbA1c follow-up | 38.79 [7.82] | 37.16 [7.42] | 39.02 [10.09] | 36.94 [8.38] |
| ∆HbA1c | 0.10 [4.82] | -0.70 [4.74] | 0.46 [6.78] | -0.37 [5.89] |
| eGFR |  |  |  |  |
| eGFR baseline | 100.9 [17.30] | 101.7 [16.62] | 98.43 [15.95] | 99.37 [15.38] |
| eGFR follow-up | 90.16 [16.97] | 92.52 [16.50] | 87.96 [16.15] | 90.23 [15.86] |
| ∆eGFR | -10.80 [9.19] | -9.32 [9.58] | -10.45 [8.97] | -9.10 [9.38] |

*Values are presented as means [SD]. SBP, systolic blood pressure; DBP, diastolic blood pressure; TC, total cholesterol; FPG, fasting plasma glucose; HbA1c, haemoglobin A1c; eGFR, estimated glomerular filtration rate; SD, standard deviation. For some factors, the change between baseline and follow-up is in the opposite direction as expected, for example, SBP and DBP. This can likely be attributed to several factors, including the healthy participant effect, the regression to the mean, the effect of medication (as participants who had hypertension at baseline were informed about this, and might thus have received medication), and finally, the effect of the pandemic.

**Supplemental Table 2. Linear regressions on the association between exposure to the pandemic, including lockdown measures, and temporal change in metabolic risk factors for those without medication at baseline, by sex***

|  | Women | | Men | | | |
| --- | --- | --- | --- | --- | --- | --- |
|  | Β [95% CI] | p-value | | Β [95% CI] | p-value | |
| No HT or DM medication |  |  | |  | |  |
| ∆SBP | 1.109 [0.097, 2.121] | .032 | | 1.698 [0.544, 2.852] | | .004 |
| ∆DBP | 1.155 [0.561, 1.750] | .000 | | 1.513 [0.754, 2.272] | | .000 |
| ∆TC | -0.032 [-0.098, 0.034] | .335 | | 0.018 [-0.056, 0.092] | | .626 |
| ∆FPG | 0.166 [0.117, 0.216] | .000 | | 0.059 [-0.001, 0.119] | | .055 |
| ∆HbA1c | -0.531 [-0.832, -0.230] | .001 | | -0.345 [-0.774, 0.085] | | .116 |
| ∆eGFR | 1.036 [0.284, 1.788] | .007 | | 1.398 [0.548, 2.248] | | .001 |
| No HT medication |  |  | |  | |  |
| ∆SBP | 1.460 [0.443, 2.478] | .005 | | 1.758 [0.619, 2.897] | | .003 |
| ∆DBP | 1.245 [0.654, 1.836] | .000 | | 1.490 [0.742, 2.239] | | .000 |
| ∆TC | -0.039 [-0.105, 0.028] | .258 | | 0.027 [-0.048, 0.101] | | .480 |
| ∆FPG | 0.140 [0.075, 0.206] | .000 | | 0.058 [-0.019, 0.136] | | .141 |
| ∆HbA1c | -0.491 [-0.829, -0.153] | .004 | | -0.368 [-0.919, 0.182] | | .190 |
| ∆eGFR | 1.091 [0.346, 1.835] | .004 | | 1.392 [0.554, 2.230] | | .001 |
| No DM medication |  |  | |  | |  |
| ∆SBP | 0.716 [-0.359, 1.792] | .192 | | 1.721 [0.604, 2.838] | | .003 |
| ∆DBP | 0.643 [0.018, 1.269] | .044 | | 1.080 [0.374, 1.786] | | .003 |
| ∆TC | -0.047 [-0.108, 0.014] | .130 | | -0.011 [-0.080, 0.059] | | .763 |
| ∆FPG | 0.139 [0.094, 0.184] | .000 | | 0.039 [-0.018, 0.096] | | .184 |
| ∆HbA1c | -0.773 [-1.046, -0.500] | .000 | | -0.679 [-1.062, -0.296] | | .001 |
| ∆eGFR | 0.955 [0.274, 1.636] | .006 | | 1.284 [0.576, 1.992] | | .000 |

*SBP, systolic blood pressure; DBP, diastolic blood pressure; TC, total cholesterol; FPG, fasting plasma glucose; HbA1c, haemoglobin A1c; eGFR, estimated glomerular filtration rate; CI, confidence intervals. The analyses were weighted on baseline measurements, age at baseline, educational level, ethnicity, occupational level and labour market participation.

**Supplemental Table 3. Linear regression on the association between exposure to the pandemic, including lockdown measures, and temporal change in metabolic risk factors, only for those assessed between July and December, by sex***

|  | Women | | Men | |
| --- | --- | --- | --- | --- |
|  | Β [95% CI] | p-value | Β [95% CI] | p-value |
| ∆SBP | 1.432 [0.111, 2.753] | .034 | 1.567 [0.207, 2.926] | .024 |
| ∆DBP | 0.941 [0.183, 1.699] | .015 | 1.014 [0.143, 1.884] | .023 |
| ∆TC | -0.047 [-0.123, 0.030] | .234 | 0.016 [-0.070, 0.103] | .712 |
| ∆FPG | 0.116 [0.035, 0.198] | .005 | 0.083 [-0.023, 0.189] | .126 |
| ∆HbA1c | -0.423 [-0.826, -0.019] | .040 | -0.192 [-0.853, 0.469] | .569 |
| ∆eGFR | 1.278 [0.459, 2.096] | .002 | 1.003 [0.092, 1.915] | .031 |

*SBP, systolic blood pressure; DBP, diastolic blood pressure; TC, total cholesterol; FPG, fasting plasma glucose; HbA1c, haemoglobin A1c; eGFR, estimated glomerular filtration rate; CI, confidence intervals. The analyses were weighted on baseline measurements, age at baseline, educational level, ethnicity, occupational level and labour market participation.

**Supplemental Table 4. Linear regressions on the association between exposure to the pandemic, including lockdown measures, and temporal change in metabolic risk factors, by sex and ethnicity***

|  | Women | | Men | |
| --- | --- | --- | --- | --- |
|  | Β [95% CI] | p-value | Β [95% CI] | p-value |
| Dutch |  |  |  |  |
| ∆SBP | 2.263 [0.578, 3.947] | .009 | 0.550 [-1.274, 2.375] | .554 |
| ∆DBP | 2.130 [1.169, 3.090] | .000 | 0.650 [-0.498, 1.799] | .267 |
| ∆TC | -0.026 [-0.136, 0.083] | .636 | 0.038 [-0.096, 0.171] | .582 |
| ∆FPG | 0.226 [0.167, 0.284] | .000 | 0.108 [-0.008, 0.223] | .067 |
| ∆HbA1c | -0.608 [-0.960, -0.256] | .001 | -0.657 [-1.280, -0.034] | .039 |
| ∆eGFR | 1.359 [0.108, 2.611] | .033 | 1.545 [0.507, 2.583] | .004 |
| South-Asian Surinamese |  |  |  |  |
| ∆SBP | 1.475 [-1.394, 4.343] | .313 | 3.023 [0.285, 5.761] | .031 |
| ∆DBP | 0.989 [-0.647, 2.624] | .236 | 1.617 [-0.233, 3.466] | .087 |
| ∆TC | -0.232 [-0.412, -0.052] | .012 | 0.031 [-0.175, 0.237] | .767 |
| ∆FPG | 0.072 [-0.102, 0.246] | .416 | 0.170 [-0.069, 0.408] | .163 |
| ∆HbA1c | -1.063 [-1.898, -0.229] | .013 | -0.077 [-1.746, 1.592] | .928 |
| ∆eGFR | 0.229 [-1.292, 1.751] | .767 | -0.119 [-2.166, 1.927] | .909 |
| African Surinamese |  |  |  |  |
| ∆SBP | -1.062 [-3.493, 1.369] | .391 | 1.497 [-1.319, 4.312] | .297 |
| ∆DBP | -1.216 [-2.685, 0.253] | .105 | 1.020 [-0.725, 2.766] | .251 |
| ∆TC | -0.111 [-0.248, 0.027] | .114 | -0.022 [-0.180, 0.136] | .782 |
| ∆FPG | 0.086 [-0.075, 0.246] | .297 | -0.034 [-0.281, 0.213] | .786 |
| ∆HbA1c | -0.317 [-1.100, 0.466] | .428 | -0.541 [-1.999, 0.918] | .467 |
| ∆eGFR | 0.612 [-0.816, 2.039] | .401 | -1.081 [-3.304, 1.143] | .340 |
| Ghanaian |  |  |  |  |
| ∆SBP | 1.247 [-2.915, 5.409] | .556 | 0.662 [-4.311, 5.635] | .793 |
| ∆DBP | 1.868 [-0.327, 4.063] | .095 | 0.569 [-2.359, 3.496] | .702 |
| ∆TC | 0.028 [-0.186, 0.242] | .797 | 0.014 [-0.234, 0.262] | .912 |
| ∆FPG | 0.266 [-0.041, 0.573] | .089 | 0.744 [0.076, 1.411] | .029 |
| ∆HbA1c | -0.717 [-2.259, 0.826] | .361 | -1.154 [-3.203, 0.895] | .268 |
| ∆eGFR | 3.015 [0.676, 5.355] | .012 | 0.898 [-2.015, 3.811] | .544 |
| Turkish |  |  |  |  |
| ∆SBP | -0.290 [-3.368, 2.788] | .853 | -0.404 [-3.439, 2.631] | .793 |
| ∆DBP | 0.590 [-1.201, 2.380] | .517 | -0.759 [-2.842, 1.323] | .474 |
| ∆TC | 0.026 [-0.166, 0.218] | .788 | -0.191 [-0.400, 0.019] | .074 |
| ∆FPG | 0.085 [-0.096, 0.266] | .358 | -0.033 [-0.223, 0.158] | .736 |
| ∆HbA1c | 0.102 [-1.118, 1.321] | .870 | -1.115 [-2.306, 0.076] | .066 |
| ∆eGFR | 0.395 [-1.369, 2.160] | .660 | 1.471 [-0.472, 3.413] | .137 |
| Moroccan |  |  |  |  |
| ∆SBP | 1.457 [-1.141, 4.055] | .271 | 1.122 [-1.320, 3.564] | .367 |
| ∆DBP | 0.465 [-0.897, 1.827] | .503 | 0.911 [-0.614, 2.436] | .241 |
| ∆TC | 0.049 [-0.088, 0.187] | .481 | -0.067 [-0.207, 0.074] | .350 |
| ∆FPG | -0.003 [-0.185, 0.179] | .976 | -0.215 [-0.402, -0.028] | .024 |
| ∆HbA1c | -0.390 [-1.188, 0.408] | .337 | -0.411 [-1.833, 1.012] | .571 |
| ∆eGFR | 1.271 [-0.414, 2.955] | .139 | 1.179 [-0.303, 2.661] | .119 |

*SBP, systolic blood pressure; DBP, diastolic blood pressure; TC, total cholesterol; FPG, fasting plasma glucose; HbA1c, haemoglobin A1c; eGFR, estimated glomerular filtration rate; CI, confidence intervals. The analyses were weighted on baseline measurements, age at baseline, educational level, occupational level and labour market participation.

**Supplemental Table 5. Linear regressions on the association between exposure to the pandemic, including lockdown measures, and temporal change in metabolic risk factors, by sex and educational level (as a proxy of socioeconomic status)***

|  | Women | | Men | |
| --- | --- | --- | --- | --- |
|  | Β [95% CI] | p-value | Β [95% CI] | p-value |
| Lower-educated |  |  |  |  |
| ∆SBP | 2.598 [0.746, 4.449] | .006 | 2.004 [0.153, 3.856] | .034 |
| ∆DBP | 1.253 [0.258, 2.248] | .014 | 0.798 [-0.205, 1.800] | .119 |
| ∆TC | -0.085 [-0.194, 0.024] | .126 | 0.002 [-0.106, 0.110] | .974 |
| ∆FPG | 0.050 [-0.082, 0.181] | .459 | -0.065 [-0.220, 0.089] | .409 |
| ∆HbA1c | -0.498 [-1.167, 0.171] | .144 | -1.123 [-1.999, -0.248] | .012 |
| ∆eGFR | 0.207 [-0.831, 1.245] | .696 | 1.111 [0.016, 2.206] | .047 |
| Intermediate-educated |  |  |  |  |
| ∆SBP | 0.828 [-1.034, 2.691] | .383 | 0.552 [-1.407, 2.512] | .580 |
| ∆DBP | 0.184 [-0.957, 1.324] | .752 | 0.971 [-0.313, 2.254] | .138 |
| ∆TC | -0.061 [-0.167, 0.045] | .260 | -0.120 [-0.243, 0.003] | .055 |
| ∆FPG | 0.130 [0.018, 0.242] | .023 | 0.033 [-0.155, 0.220] | .733 |
| ∆HbA1c | -0.945 [-1.564, -0.326] | .003 | -0.198 [-1.112, 0.717] | .671 |
| ∆eGFR | 2.014 [0.707, 3.321] | .003 | 0.225 [-1.173, 1.623] | .752 |
| Higher-educated |  |  |  |  |
| ∆SBP | 0.209 [-1.406, 1.825] | .799 | 1.650 [-0.009, 3.309] | .051 |
| ∆DBP | 1.207 [0.058, 1.995] | .038 | 1.033 [-0.066, 2.132] | .065 |
| ∆TC | 0.014 [-0.076, 0.104] | .766 | 0.050 [-0.059, 0.158] | .369 |
| ∆FPG | 0.112 [0.042, 0.183] | .002 | 0.073 [-0.014, 0.159] | .100 |
| ∆HbA1c | -0.896 [-1.287, -0.505] | .000 | -0.784 [-1.523, -0.046] | .038 |
| ∆eGFR | 0.987 [-0.050, 2.025] | .062 | 1.360 [0.300, 2.420] | .012 |

*SBP, systolic blood pressure; DBP, diastolic blood pressure; TC, total cholesterol; FPG, fasting plasma glucose; HbA1c, haemoglobin A1c; eGFR, estimated glomerular filtration rate; CI, confidence intervals. The analyses were weighted on baseline measurements, age at baseline, ethnicity, occupational level and labour market participation.

**Supplemental Table 6. Mediators at baseline, follow-up and change over time by experimental group and sex**

|  | Women | | Men | |
| --- | --- | --- | --- | --- |
|  | Control | Exposed | Control | Exposed |
| BMI |  |  |  |  |
| BMI baseline | 27.0 [5.31] | 26.3 [5.42] | 26.0 [3.85] | 25.5 [3.62] |
| BMI follow-up | 27.6 [5.43] | 27.3 [5.68] | 26.5 [3.98] | 26.2 [3.90] |
| ∆BMI | 0.694 [2.17] | 0.956 [2.51] | 0.422 [1.79] | 0.627 [1.73] |
| PHQ-9 |  |  |  |  |
| PHQ-9 baseline | 4.35 [4.58] | 4.56 [4.62] | 3.24 [4.06] | 3.15 [3.74] |
| PHQ-9 follow-up | 4.65 [4.57] | 4.70 [4.26] | 3.40 [3.97] | 3.21 [3.44] |
| ∆PHQ | 0.320 [4.58] | 0.149 [4.56] | 0.155 [3.86] | 0.071 [3.64] |
| ∆Smoking (%) |  |  |  |  |
| No change | 2613 [91.7] | 949 [90.6] | 1940 [86.8] | 675 [83.2] |
| Quit | 152 [5.3] | 78 [7.4] | 172 [7.7] | 70 [8.6] |
| Started | 84 [3.0] | 21 [2.0] | 122 [5.5] | 66 [8.1] |
| ∆Alcohol (%) |  |  |  |  |
| Less | 178 [7.9] | 160 [15.3] | 177 [10.2] | 135 [16.7] |
| Equal | 1971 [87.9] | 831 [79.5] | 1460 [84.0] | 619 [76.7] |
| More | 94 [4.2] | 55 [5.3] | 101 [5.8] | 53 [6.6] |
| NLE at follow-up (%) | 1901 [66.6] | 698 [66.5] | 1408 [63.2] | 496 [61.2] |

*BMI and PHQ-9 values are presented as means [SD], while smoking, alcohol use and NLE were presented as frequencies [percentages]. BMI, body mass index; PHQ, patient health questionnaire; NLE, negative life event; SD, standard deviation.

**Supplemental Table 7. Linear regressions on the association between exposure to the pandemic, including lockdown measures, and temporal change in metabolic risk factors by sex for early and late post-lockdown period***

|  | Women | | | | Men | | | |
| --- | --- | --- | --- | --- | --- | --- | --- | --- |
|  | Early post-lockdown period | | Late post-lockdown period | | Early post-lockdown period | | Late post-lockdown period | |
|  | Β [95% CI] | p-value | Β [95% CI] | p-value | Β [95% CI] | p-value | Β [95% CI] | p-value |
| ∆SBP | 0.909 [-0.468, 2.286] | .196 | 2.106 [0.805, 3.406] | .002 | 0.965 [-0.528, 2.457] | .205 | 2.251 [0.964, 3.538] | .001 |
| ∆DBP | 0.294 [-0.456, 1.044] | .442 | 1.310 [0.533, 2.087] | .001 | 0.417 [-0.531, 1.366] | .388 | 1.428 [0.618, 2.238] | .001 |
| ∆TC | -0.063 [-0.142, 0.015] | .114 | -0.044 [-0.123, 0.035] | .273 | -0.081 [-0.176, 0.013] | .092 | 0.085 [0.000, 0.171] | .049 |
| ∆FPG | 0.081 [0.006, 0.156] | .034 | 0.153 [0.069, 0.236] | .000 | 0.010 [-0.089, 0.108] | .848 | 0.151 [0.034, 0.268] | .011 |
| ∆HbA1c | -1.134 [-1.519, -0.749] | .000 | -0.264 [-0.670, 0.142] | .202 | -1.101 [-1.763, -0.440] | .001 | 0.143 [-0.538, 0.823] | .681 |
| ∆eGFR | 1.246 [0.434, 2.057] | .003 | 1.390 [0.560, 2.220] | .001 | 0.868 [-0.088, 1.825] | .075 | 1.361 [0.470, 2.251] | .003 |

*SBP, systolic blood pressure; DBP, diastolic blood pressure; TC, total cholesterol; FPG, fasting plasma glucose; HbA1c, haemoglobin A1c; eGFR, estimated glomerular filtration rate; CI, confidence intervals. The analyses were weighted on baseline measurements, age at baseline, educational level, ethnicity, occupational level and labour market participation.
